# Projected burden of hypertension-associated cardiovascular disease in people living with HIV versus HIV-negative adults in Eswatini

**DOI:** 10.64898/2026.07.17.26358301

**Authors:** Masabho P. Milali, Daniel T. Citron, Kasturi Bhamidipati, Nao Yamamoto, Afia Osei-Ntansah, Ingrida Platais, Gianna Ferrara, Cebisile Ngcamphalala, Sotja G. Dlamini, Ntombifuthi Ginindza, Anna Bershteyn

## Abstract

**Background:** Having achieved the UNAIDS 95–95–95 targets, Eswatini faces a growing burden of non-communicable diseases which are major contributors to morbidity and mortality. Hypertension-associated cardiovascular disease (CVD) is rising among people living with HIV (PLHIV) as survival improves and metabolic risks – including those linked to dolutegravir (DTG) – increase. We projected CVD burden among PLHIV and HIV-negative adults (PLWHIV) through 2045 to inform integrated HIV–CVD planning.

**Methods:** EMOD-HIV, an agent-based model calibrated to Eswatini’s epidemic, generated HIV prevalence trajectories. These were combined with age-standardized Global Burden of Disease CVD estimates and published relative risks (RRs) to produce HIV-stratified CVD projections. CVD burden trajectories were then projected through 2045 using a logistic generalized additive model with Monte Carlo uncertainty quantification. Five scenarios were evaluated to assess how different assumptions about RR of CVD among PLHIV versus PLWHIV affect projected burden: (1) CVD prevalence under a constant RR; (2) CVD mortality under a constant RR; (3) HTN-attributable CVD mortality under a constant RR; (4) HTN-attributable CVD mortality under a post-DTG RR increase following Eswatini’s 2021 dolutegravir rollout; and (5) HTN-attributable CVD mortality under a gradual RR increase from 2010–2045 reflecting cumulative metabolic and demographic shifts.

**Results:** PLHIV consistently exhibited higher CVD burden than HIV-negative adults. Scenario 1: CVD prevalence was 11.0% (95% UI: 9.0-13.5%) among PLHIV versus 6.8% (6.0-7.8%) in 2025, stable through 2045. Scenario 2: CVD mortality rate was 0.60% (0.47-0.76%) versus 0.37% (0.31-0.45%) in 2025, declining modestly through 2045 with consistent excess. Scenario 3: HTN-attributable mortality was 73% (70-76%) versus 64% (61-66%) in women and 61% (58-64%) versus 58% (55-60%) in men, stable through 2045. Scenario 4: Following DTG rollout, mortality rose from 73% to 85% in women and 61% to 70% in men by 2022, remaining stable thereafter. Scenario 5: By 2045, mortality reached 85% (82-88%) in women and 67% (64-70%) in men with HIV, versus 60% (57 – 63%) and 55% (53 – 57%) in HIV-negative adults.

**Conclusions:** While excess CVD burden among PLHIV is projected to persist even under stable risk conditions, ART-related metabolic trajectories – particularly those linked to DTG – may drive substantial widening of this gap through 2045. HTN-attributable CVD mortality is particularly elevated among women with HIV. Strengthening integrated HIV–NCD services, including blood pressure screening, risk-based therapy, and sex-specific DTG counseling, will be essential to sustain long-term health gains.

## Introduction

Cardiovascular disease (CVD) has emerged as a leading cause of morbidity and mortality in sub-Saharan Africa, accounting for nearly one-third of all non-communicable disease (NCD) deaths (1,2). In Eswatini, CVD risk is compounded by the world’s highest HIV prevalence—approximately 25% among adults aged 15–49 years (3–5), and the country’s rapid epidemiologic transition toward NCDs (6). As people living with HIV (PLHIV) experience longer survival under antiretroviral therapy (ART), comorbidities such as hypertension, diabetes, and dyslipidemia have become increasingly common (3,7). In settings with high ART coverage and viral suppression, these chronic conditions contribute an increasing share of morbidity and mortality among adults with HIV (8).

Growing evidence links both HIV infection and ART exposure to elevated cardiometabolic risk through mechanisms including chronic immune activation, endothelial dysfunction, and metabolic effects of modern antiretroviral regimens (7,9–12). Notably, dolutegravir (DTG)–based ART, now first-line in Eswatini’s national program, has been associated with accelerated weight gain and increased risk of hypertension, particularly among women (9,10,13,14). As the country expands DTG use, these metabolic shifts may interact with population aging and rising background hypertension prevalence, amplifying the CVD burden among PLHIV.

Eswatini has made substantial progress in integrating NCD services within HIV care, including routine blood pressure screening, diabetes and lipid testing, and lifestyle counseling (5). However, the long-term implications of ART-related metabolic changes for national CVD trends among PLHIV in Eswatini remain unclear. Few studies in Eswatini address DTG-associated weight gain (3,14,16) and implementation of CVD risk screening within HIV services. (15,17) To date, no studies have attempted to quantify how evolving ART patterns, population aging, and secular trends in adiposity and hypertension will jointly shape future CVD burden over the next decades.

To address this gap, we developed demographic and HIV epidemiological projections using the agent-based Epidemiological MODeling–HIV (EMOD-HIV) platform (18,19), calibrated to Eswatini’s HIV epidemic, and integrated it with hypertension and CVD parameters derived from the Global Burden of Disease (GBD) (20) and WHO STEPwise (STEPS) surveys (21). We simulated CVD- and hypertension-attributable mortality through 2045 under alternative relative-risk (RR) scenarios reflecting key epidemiologic transitions, including ART-related metabolic effects, survival-driven population aging among PLHIV, and secular increases in adiposity and hypertension. We hypothesized that these interacting transitions would lead to a persistently higher—and potentially widening—CVD burden among people living with HIV compared with HIV-negative adults over time. The aim of this study was to project and compare long-term CVD burden by HIV status under plausible future risk trajectories to inform integrated HIV–NCD planning in Eswatini.

## Methods

### Ethics approval and consent to participate

This study did not involve human participants, human specimens, or identifiable individual-level data. The analysis was based on mathematical modeling using publicly available, aggregated data from the Global Burden of Disease (GBD) study, World Health Organization STEPwise (STEPS) surveys, published literature, and outputs from a previously developed and calibrated EMOD-HIV simulation model. Therefore, institutional ethics approval and informed consent were not required.

### Study overview

This modeling study used the Epidemiological MODeling–HIV (EMOD-HIV) agent-based model (18,19) to forecast the burden of hypertension (HTN)-associated cardiovascular disease (CVD) in Eswatini over a 20-year horizon (2025 to 2045). HTN-associated CVD was defined as acute myocardial infarction (AMI/heart attack), stroke (ischemic or hemorrhagic), or heart failure. EMOD-HIV was first calibrated to replicate Eswatini’s population structure and HIV epidemic dynamics using data from the 2006–7 Demographic and Health Survey (DHS) (22,23) and the Swaziland HIV Incidence Measurement Surveys (SHIMS) conducted 2011, 2016–17, and 2021 (24,25). Because EMOD-HIV does not natively simulate non-communicable diseases, CVD parameters were incorporated in a post-processing step outside the model, using age-standardized estimates obtained directly from the Global Burden of Disease (GBD) study (20). Three related but conceptually distinct outcomes were derived from these GBD datasets: (1) CVD prevalence – the proportion of the total adult population living with CVD in a given year (denominator: total population); (2) CVD mortality rate – the annual risk of dying from CVD, expressed as deaths per 100,000 person-years (denominator: total population); and (3) the proportion of CVD deaths attributable to elevated systolic blood pressure – the hypertension-mediated share of CVD mortality (denominator: total CVD deaths, not total population). These metrics are complementary but not directly comparable, as they express burden relative to different reference groups. These age-standardized CVD prevalence estimates were then combined with age- and sex-specific HIV prevalence trajectories generated by EMOD-HIV to reconstruct HIV-negative and HIV-positive CVD prevalence time series for the Eswatini adult population. Population-level prevalence was obtained by weighting HIV-stratified prevalence by the corresponding HIV prevalence in each year. Projections were stratified by HIV status, enabling direct comparisons between people living with HIV (PLHIV) and HIV-negative individuals.

### Model description

EMOD-HIV is an agent-based simulation framework developed by the Institute for Disease Modeling at the Gates Foundation and made publicly available as open-source software (19,26). It captures both heterosexual and vertical transmission, HIV disease progression, and transitions along the HIV prevention and care continuum (19,26). Sexual relationships are modeled explicitly (long-term, informal, short-term, and transactional), forming and dissolving dynamically based on demographic and behavioral characteristics. Transmission risks are modeled per coital act and at birth, with modifications reflecting prevention and treatment uptake that alter infectiousness or susceptibility (19,26). EMOD has been extensively validated and applied in multiple African settings to assess HIV prevention and treatment strategies (27–29).

### Model calibration

EMOD-HIV was calibrated to Eswatini using SHIMS (24) and DHS (22) data. Calibration employed parallel simultaneous perturbation optimization, a stochastic gradient descent algorithm adapted for parallel computing (30,31), to tune uncertain parameters related to epidemic initialization and sexual behavior. Targets included population age and sex structure (from census data), age- and sex-specific HIV prevalence and incidence (from SHIMS and DHS), and HIV service coverage (from national health agencies). The timeline of HIV care was parameterized based on SHIMS and DHS data, tracking the expanded distribution of ART, voluntary medical male circumcision (VMMC), and antenatal care. The distribution of PrEP was parameterized using data from PrEPWatch (32). We generated 250 calibrated epidemic trajectories using a likelihood-weighted roulette sampling approach (33), representing alternative but plausible epidemic histories consistent with observed data. These trajectories formed the basis for estimating age- and sex-specific HIV prevalence over time. Model performance was validated against SHIMS data from 2007, 2011, 2016, and 2021 (Supplementary Figures S1–S2).

### Integration of CVD and HTN-associated CVD parameters

This section describes the data sources, estimation and forecasting approaches for CVD prevalence, CVD mortality, and HTN-attributable CVD deaths, and the propagation of uncertainty from data sources to outcomes.

#### Data sources

Three age-standardized inputs (CVD prevalence, CVD mortality rates, and HTN-attributable CVD deaths) were obtained from the GBD (20), covering annual estimates from 2010 – 2021. CVD prevalence was extracted as age-standardized percent, expressed as the proportion of the total population living with CVD (Supplementary Figure S3). CVD mortality rates were extracted as age-standardized rates per 100,000 person-years and converted to annual CVD death probabilities expressed as a percentage of the total population (÷ 100,000 × 100) (Supplementary Figure S4). HTN-attributable CVD deaths were extracted as the age-standardized proportion of CVD deaths attributable to high systolic blood pressure; the denominator for this metric is total CVD deaths within each sex stratum — not the total population — and is therefore not directly comparable to CVD prevalence or mortality rate estimates (Supplementary Figure S5). For all three inputs, no additional statistical manipulation was applied beyond deriving standard deviations from the reported GBD 95% uncertainty intervals using SD = (upper − lower) ÷ 3.92, which were propagated through Monte Carlo simulation. WHO STEPwise (STEPS) risk factor surveys (21) conducted in 2014 and 2024 were used for contextual validation of HTN prevalence trends but were not used as primary inputs to the modeling because they provide only two time points and were consistent with GBD estimates. HIV prevalence trajectories, stratified by age and sex, were derived from calibrated EMOD-HIV model outputs.

Relative risk (RR) estimates comparing PLHIV with HIV-negative individuals were extracted from published literature (1,12,34,35) and are summarized in Supplementary Tables S1 and S2. For HTN-associated CVD, studies commonly reported hazard ratios (HRs) by HIV status rather than direct comparative relative risks (RRs) (13). We converted HRs to approximate RRs using Equation 1 (36). Although this formula was originally derived for odds ratio-to-RR conversion, it is appropriate here because when annual baseline risks are small — as is the case in this study (≤0.4% per 100,000 population in HIV-negative adults; Supplementary Table S1) — HRs, odds ratios, and RRs are numerically similar, allowing the OR-based correction to be applied to HRs with minimal error (37).

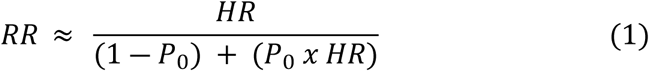

where *P*_0_ represents the baseline risk of the outcome among HIV-negative adults, estimated from GBD 2021 age-standardized CVD mortality rates for Eswatini. Because GBD reports rates per 100,000 population rather than cumulative risks, *P*_0_ was derived under the exponential survival assumption using Equation 2 (38):

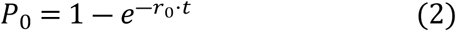

where *r*_0_ is the GBD age-standardized HTN-attributable CVD mortality rate (per 100,000 population, converted to a proportion) used solely to derive the baseline risk *P*_0_ required for the HR-to-RR conversion; general (unstratified) CVD mortality is modeled separately as a distinct forecast outcome. *t* is the follow-up duration, aligned with the period over which the HRs were reported. A step-by-step description of the HR-to-RR conversion is provided in the Supplementary methods (Conversion of HRs to RRs), with intermediate quantities summarized in Supplementary Table S1. Because published studies (34) reported HRs separately for HIV-positive and HIV-negative adults, each expressed relative to a common external reference group rather than relative to each other, neither HR directly quantifies the excess CVD risk in PLHIV compared with HIV-negative adults. We therefore first applied Equation 1 to convert each group-specific HR into an approximate cumulative risk, then obtained the RR of HTN-associated CVD comparing PLHIV with HIV-negative adults as the ratio of these two estimated risks (Supplementary Table S2). For general CVD, RRs directly comparing PLHIV with HIV-negative adults were available from prior studies (12,35), reflecting a broader evidence base for overall CVD outcomes; by contrast, HTN-associated CVD studies typically reported hazard ratios stratified by HIV status rather than direct comparative RRs, necessitating the conversion approach described above.

#### Uncertainty in data sources

GBD estimates include uncertainty from data limitations and model specification and are reported with 95% uncertainty intervals (20). STEPS survey estimates reflect sampling variability and measurement error and are reported with survey-based confidence intervals (21). Published relative risk estimates include reported 95% confidence intervals(12,34,35). These uncertainties were propagated through the analysis using probabilistic sensitivity analysis, as described below.

#### Estimating and Forecasting CVD Prevalence

We estimated and projected cardiovascular disease (CVD) prevalence among PLHIV and HIV-negative adults through 2045 using a multi-step modeling framework (Supplementary Methods). General population age-standardized CVD prevalence from the GBD 2010–2021 dataset (Supplementary Figure S3) was decomposed into HIV-negative and PLHIV subgroups using HIV prevalence trajectories from EMOD-HIV and relative risk (RR) estimates from published literature (Supplementary Table S2) comparing PLHIV with HIV-negative adults. CVD prevalence among HIV-negative adults was calculated as:

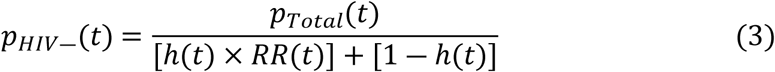

where *p*_*Total*_(*t*) denotes the general population CVD prevalence, ℎ(*t*) is the HIV prevalence in year *t*, and *RR*(*t*) is the relative risk of CVD among PLHIV compared with HIV-negative adults. Corresponding CVD prevalence among PLHIV was then obtained by applying the same relative risk to the decomposed HIV-negative estimates:

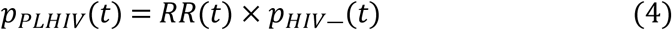

These reconstructed time series (2010–2021) represent the empirical baseline derived from GBD, EMOD-HIV, and literature inputs. They served as the basis for forecasting future trends, with uncertainty propagated through Equations (3) and (4) as described in the Uncertainty Analysis section.

To project CVD prevalence beyond 2021, we fitted a generalized additive model (GAM) (39,40) with a logistic link and penalized splines (41), to the HIV-negative prevalence series. The logistic link constrains predictions between 0 and 1, while penalized splines capture smooth, nonlinear temporal trends. The model was specified as:

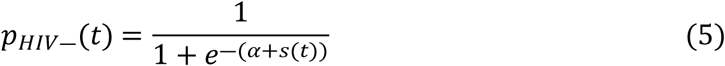

where *p*_*HIV*―_(*t*) is the predicted HIV-negative CVD prevalence in year *t*, *α* is the intercept, and *s*(*t*) is a smooth function of time estimated using penalized splines. The fitted model was then extrapolated from 2022 to 2045, assuming that long-term changes continue smoothly along the observed temporal trajectory.

Forecasted CVD prevalence among PLHIV was subsequently derived by applying the scenario-specific relative risk *RR*(*t*) to the corresponding HIV-negative baseline for each year, maintaining internal consistency across scenarios and time periods:

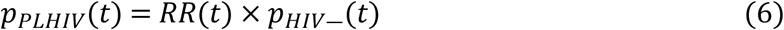

Logistic regression (42,43) with linear time was used as a sensitivity analysis.

#### Estimating and Forecasting CVD Mortality

Estimation and forecasting of CVD mortality followed the same modeling framework used for CVD prevalence, differing only in the choice of input data. Age-standardized CVD mortality rates (deaths per 100,000 person-years; denominator: total population) were obtained from the GBD study and converted to annual CVD death risks (probability of dying from CVD in a given year, expressed as a proportion of the total population) using an exponential survival assumption. These risks (Supplementary Figure S4) were then decomposed into HIV-negative and PLHIV strata using HIV prevalence trajectories from EMOD-HIV and scenario-specific relative risks. Reconstructed HIV-stratified CVD mortality time series for 2010–2021 served as the empirical baseline for forecasting. Future trends were projected by fitting generalized additive models with a logistic link to the HIV-negative mortality series and extrapolating through 2045, with mortality among PLHIV derived by applying the corresponding relative risks. Uncertainty was propagated analogously to the CVD prevalence analysis, as described in the Uncertainty Analysis section.

#### Estimating and Forecasting Proportion of CVD deaths Attributable to Hypertension

Forecasting of CVD deaths attributable to hypertension used the same approach, with inputs drawn from GBD estimates of HTN-attributable CVD mortality (Supplementary Figure S5). Age-standardized proportions of CVD deaths attributable to elevated systolic blood pressure (denominator: total CVD deaths within each HIV-status and sex stratum, not the total population) were reconstructed for HIV-negative and PLHIV populations using HIV prevalence trajectories and relative risk scenarios. These reconstructed series formed the baseline for projecting future trends through 2045 using generalized additive models, with scenario-specific relative risks governing divergence between PLHIV and HIV-negative adults. Uncertainty was propagated from GBD attribution estimates, HIV prevalence trajectories, and relative risks using the same probabilistic framework applied in the prevalence and mortality analyses.

#### Propagation of Uncertainty from Data Sources to Outcomes

Uncertainty in the HIV-negative forecasts was captured by obtaining 1,000 draws from the estimated covariance structure of the GAM penalized spline coefficients (assumed to follow a multivariate normal distribution) and conducting independent fits of the GAM for each draw, propagating variance from both the spline fit and smoothing penalty through the joint covariance matrix of the penalized spline coefficients, which preserves their non-independence. For PLHIV, uncertainty reflected both the GAM-derived variability in the HIV-negative baseline and uncertainty in RRs, which was incorporated by treating published point estimates and 95% confidence intervals as log-normal probability distributions (Supplementary Table S2) (44). For each parametric Monte Carlo simulation (45) iteration, RR values were sampled and applied to HIV-negative prevalence to generate corresponding estimates for PLHIV. A total of 1000 iterations yielded distributions of possible trajectories; the mean was reported as the central estimate, with 95% uncertainty intervals from the 2.5th–97.5th percentiles.

#### Scenarios

Five scenarios were explored. Scenario 1 forecasted general CVD burden among PLHIV and HIV-negative individuals using a published RR of 1.6 (1.3-2.0) (35), applied to age-standardized CVD prevalence (proportion of the total population living with CVD; denominator: total population). This scenario establishes the baseline magnitude of HIV-associated excess CVD prevalence under the assumption that the RR remains constant over the projection period. Scenario 2 forecasted general CVD mortality risk using the same RR of 1.6 (1.3-2.0) (31), applied to age-standardized CVD mortality rates converted to annual CVD death probabilities expressed as a percentage of the total population (÷ 100,000 × 100; denominator: total population). This scenario establishes the baseline magnitude of HIV-associated excess CVD mortality under the same constant-RR assumption, providing a mortality analogue to Scenario 1. Scenarios 3-5 focused on HTN-associated CVD, which accounts for approximately 55-65% of CVD cases globally and ∼60% of CVD deaths in Eswatini (15,46). Sex-specific RRs were used in Scenarios 3-5 (Supplementary Table S3).

In scenario 3, RRs remained unchanged throughout the projection period – representing a counterfactual of stable relative risk – using estimates from Siddiqui et al. converted using Equation 1 and summarized in Supplementary Table S2. In scenario 4, risks were stable from 2010 through 2021, then increased in 2022 following Eswatini’s national DTG rollout in 2021 (16,47), and remained constant thereafter (women 1.4 [1.25–1.50], men 1.2 [1.10–1.30], both 1.29 [1.18–1.40]). The increase reflects DTG-associated weight gain, more pronounced among women, emerging with an approximate one-year lag (9–11). Relative-risk increases of approximately 18% in women and 14% in men were derived by translating DTG-related weight gain into BMI changes using mean adult heights from WHO STEPS Eswatini surveys (15,48), then applying published BMI-CVD risk associations (10,49,50), with detailed calculations provided in the Supplementary Information. In scenario 5, risks rose progressively from 2010, anchored to the lowest early-ART-era estimates (51) (women 1.05 [1.00–1.10], men 1.05 [1.00–1.10], both 1.1 [1.05–1.15]) and reaching target RRs by 2045 (women 1.43 [1.30–1.55], men 1.25 [1.15–1.35], both 1.33 [1.22–1.45]), reflecting the multiplicative influence of DTG-associated metabolic changes, survival-driven aging of the PLHIV population as ART scale-up continues to extend life expectancy and shift age structure (3,15), and secular increases in adiposity and hypertension across the SADC region (1,15,52), with full decomposition provided in Supplementary Note S3.

### Sensitivity analyses

Sensitivity analyses were conducted to assess the robustness of projected CVD outcomes to key modeling assumptions. First, sensitivity to the choice of extrapolation model for HIV-negative CVD outcomes was evaluated by fitting a logistic regression with a linear time trend as an alternative to the generalized additive model (GAM) and comparing resulting projections across all five scenarios; this also assessed whether the declining trend observed in HIV-negative CVD under the GAM - attributable to the downward trajectory in GBD-derived inputs over 2010-2021 - was robust to alternative model specifications. Second, uncertainty in relative risk (RR) assumptions was explored probabilistically through Monte Carlo sampling of RR distributions across all scenarios, rather than fixed point estimates. Published RR point estimates and 95% confidence intervals were treated as log-normal probability distributions (Supplementary Table S2), from which RR values were sampled and applied to HIV-negative prevalence during each simulation iteration. This approach assessed sensitivity to the magnitude of HIV- and ART-associated excess CVD risk in both the general CVD scenarios (Scenarios 1 and 2) and the HTN-associated CVD scenarios (Scenarios 3–5). Finally, projections were evaluated over alternative analytic horizons (5- and 10-year periods) in addition to the full 2010–2045 horizon to assess the stability of conclusions over shorter, policy-relevant time frames. Uncertainty in HIV prevalence trajectories from EMOD-HIV was not formally propagated through the CVD decomposition and is acknowledged as a limitation.

#### Model implementation

The overall analytical workflow is shown in Supplementary Figure S3. All analyses were conducted in Python 3. EMOD-HIV simulations generated age-and sex-stratified HIV prevalence trajectories, which were used to decompose GBD-derived CVD estimates into PLHIV and HIV-negative subgroups using the RR decomposition framework described above. Three GBD inputs were used: age-standardized CVD prevalence (Scenario 1), age-standardized CVD mortality rates converted to annual death probabilities (Scenario 2), and the age-standardized proportion of CVD deaths attributable to high systolic blood pressure (Scenarios 3-5). For each input, a logistic generalized additive model (GAM) was fitted to the 2010–2021 GBD time series and extrapolated to 2045. Scenarios 1 and 2 were analyzed using both sexes combined, whereas Scenarios 3–5 used sex-stratified analyses with sex-specific relative risks. Uncertainty was propagated through Monte Carlo simulation (N = 1,000 draws) by sampling RR values from log-normal distributions defined by published 95% confidence intervals, refitting the GAM for each draw, and computing 95% uncertainty intervals from the resulting distribution of projected outcomes.

The conceptual CVD state-transition structure underlying these projections is illustrated in Figure 1. Adults were stratified by age, sex, and HIV status and could transition from alive without CVD to alive with CVD, with hypertension modeled as a risk factor modifying the transition to CVD. Individuals in the alive with CVD state could subsequently transition to CVD-related mortality, while non-CVD mortality was included as a competing risk from both alive states. HIV was not represented as a separate disease state; instead, the same model structure was applied to PLHIV and HIV-negative adults, with HIV modifying transition rates.

**Figure 1.**
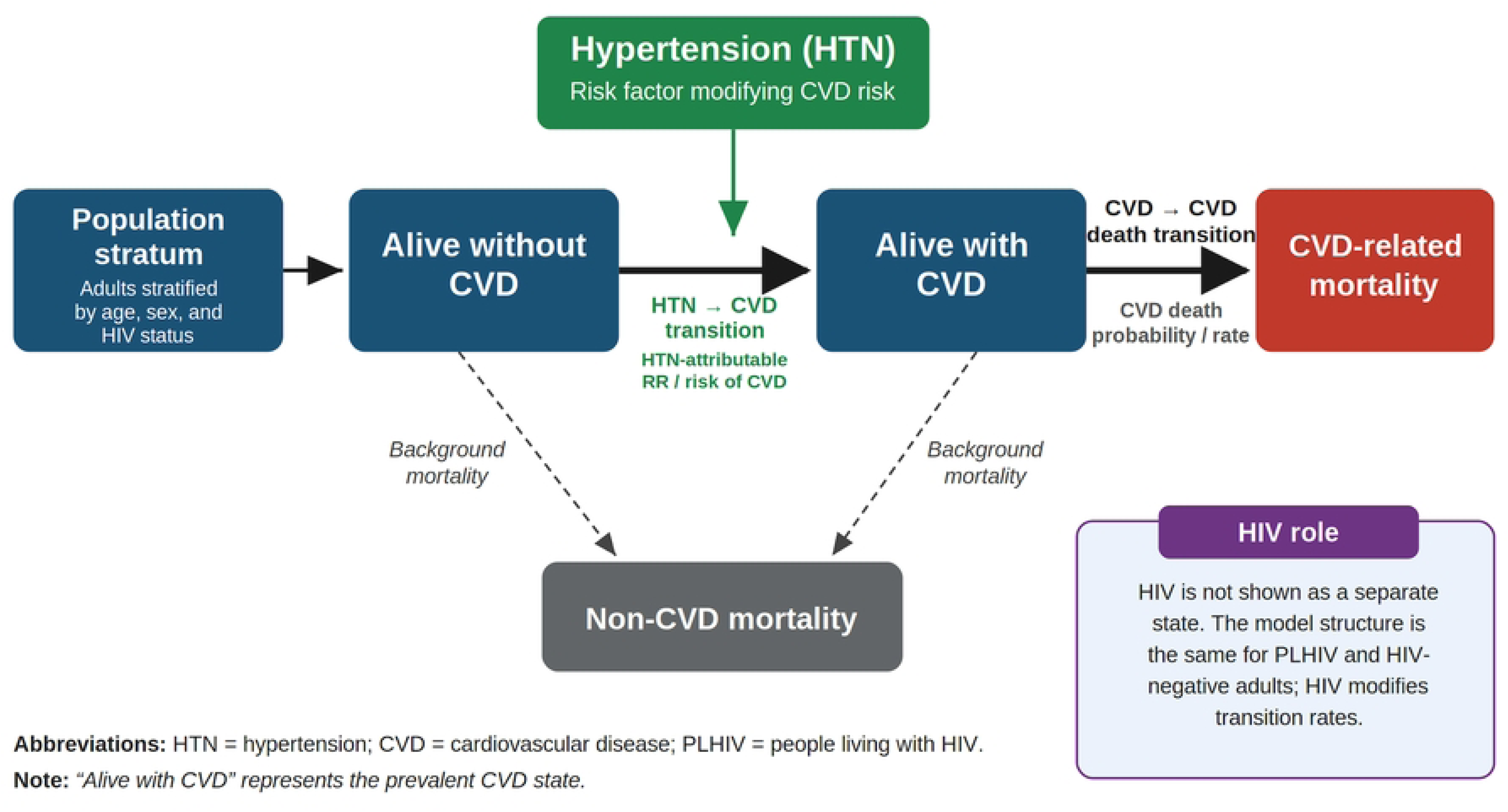
Conceptual state-transition model of CVD progression and mortality. Adults stratified by age, sex, and HIV status transition from alive without CVD to alive with CVD, and from alive with CVD to CVD-related mortality. Hypertension modifies transition to CVD, whereas non-CVD mortality acts as a competing risk from both alive states. HIV is modeled as a modifier of transition rates rather than as a separate state.

## Results

Across all scenarios, PLHIV exhibited higher CVD prevalence and a greater proportion of CVD deaths attributable to hypertension than HIV-negative adults, with the highest burden estimated among women. Consistent with the analytic framework described in the Methods, Scenario 1 characterizes baseline excess CVD burden using age-standardized prevalence (proportion of the total population living with CVD; denominator: total population) prevalence. Scenario 2 reports the general CVD mortality rate (annual probability of dying from CVD; denominator: total population). Scenarios 3–5 report the proportion of CVD deaths attributable to elevated systolic blood pressure (denominator: total CVD deaths within each HIV-status and sex stratum, not the total population), reflecting hypertension-mediated pathways rather than a population-level mortality rate.

### Scenario 1: General cardiovascular disease (constant RR)

In 2025, the model estimated an age-standardized CVD prevalence of 11.0% (95% UI: 9.0–13.5%) among PLHIV and 6.8% (95% UI: 6.0–7.8%) among HIV-negative adults. Prevalence remained stable through 2045 (PLHIV: 10.9–11.2%; HIV-negative: 6.7–6.9%), maintaining an approximately 4.2 percentage-point difference between HIV-positive and HIV-negative adults, with non-overlapping uncertainty intervals (Figure 2).

**Figure 2.**
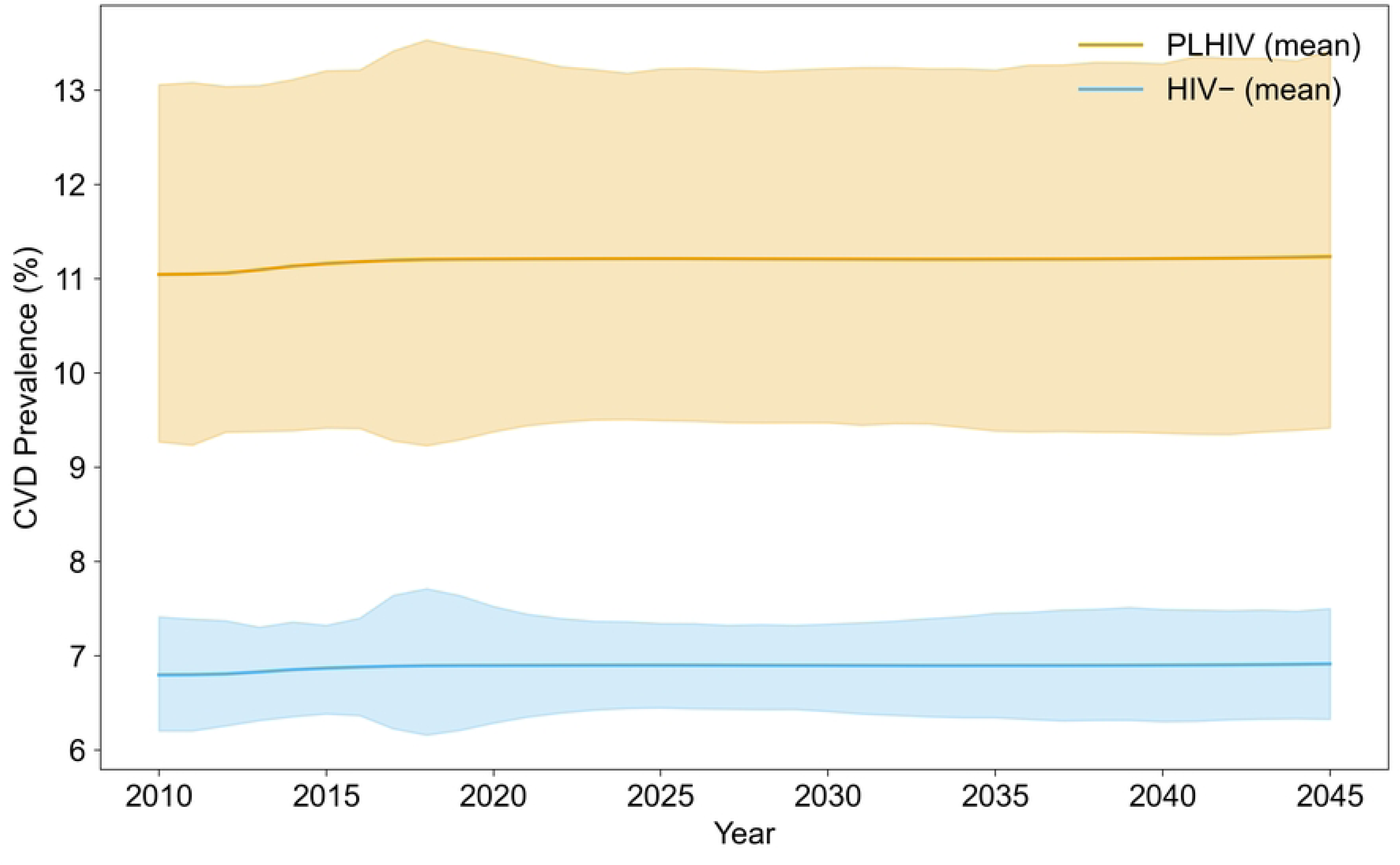
Modeled cardiovascular disease (CVD) prevalence by HIV status. Age-standardized CVD prevalence (%) with 95% uncertainty intervals under a constant relative-risk (RR = 1.6[1.3-2.0]) assumption. Projections show persistently higher CVD prevalence among PLHIV (11.0%, 95% UI 9.0-13.5%) compared with HIV-negative adults (6.8%, 95% UI 6.0-7.8%), with little change over time.

### Scenario 2: General CVD mortality (constant RR)

In 2025, the model estimated an age-standardized CVD mortality rate of 0.60% (95% UI: 0.47–0.76%) among PLHIV and 0.37% (95% UI: 0.31–0.45%) among HIV-negative adults. CVD mortality risk declined modestly through 2045, reflecting the downward trend in GBD-derived mortality inputs, reaching an estimated 0.54% (95% UI: 0.36–0.75%) among PLHIV and 0.33% (95% UI: 0.24–0.46%) among HIV-negative adults. PLHIV consistently exhibited higher CVD mortality risk than HIV-negative adults throughout the projection horizon, with 0.23 percentage points in 2025, narrowing slightly to 0.21 percentage points by 2045, with largely non-overlapping uncertainty intervals (Figure 3).

**Figure 3.**
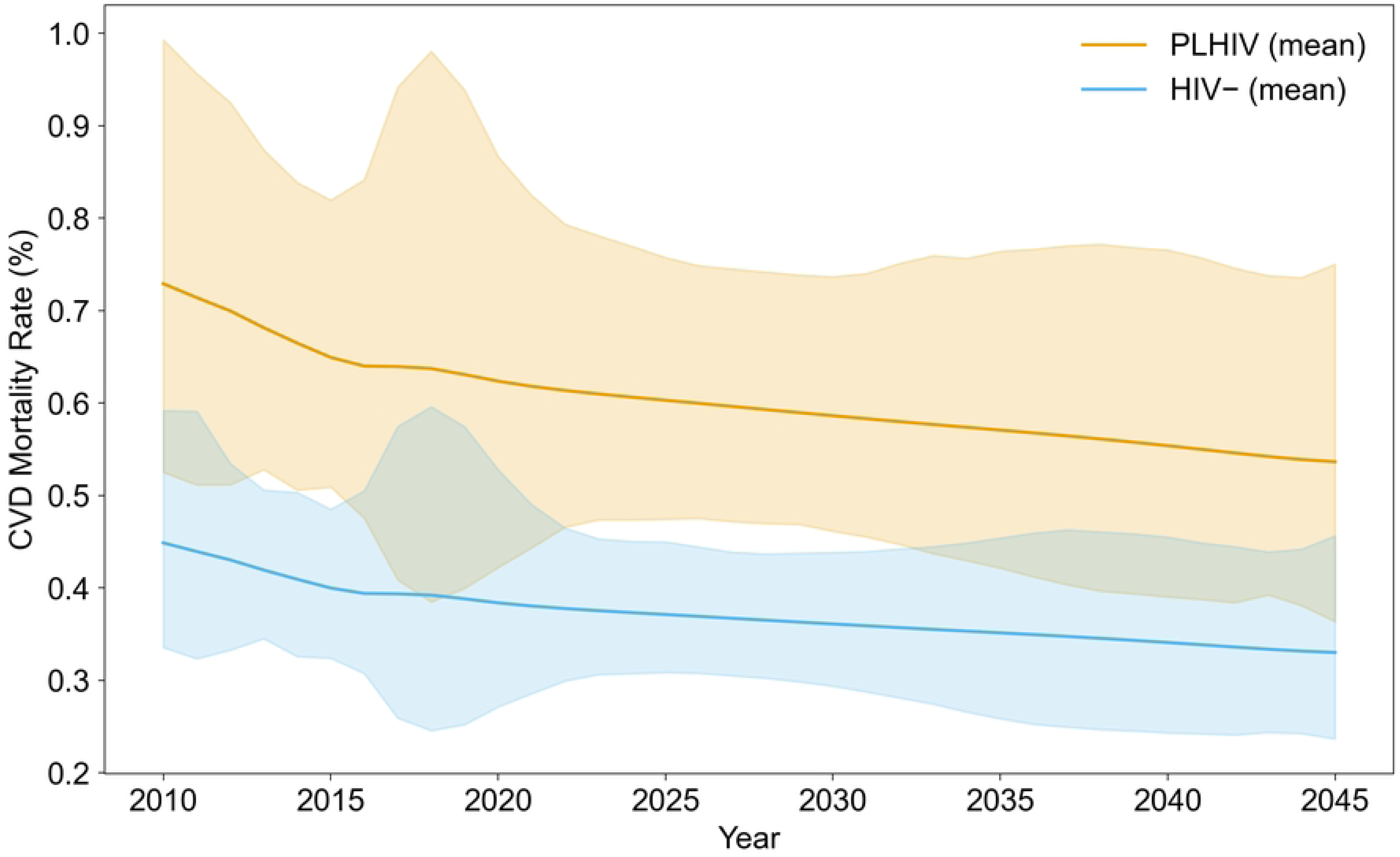
Modeled general CVD mortality risk by HIV status. Age-standardized annual probability of dying from CVD (%) with 95% uncertainty intervals under a constant relative-risk (RR = 1.6 [1.3–2.0]) assumption. PLHIV consistently exhibited higher CVD mortality risk than HIV-negative adults (0.60% vs 0.37% in 2025), with a modest declining trend reflecting GBD-derived mortality inputs and largely non-overlapping mean estimates throughout the projection horizon.

### Scenario 3: Hypertension-associated CVD (constant RR)

Under a constant relative risk for hypertension-associated CVD, the proportion of CVD deaths attributable to elevated systolic blood pressure (SBP) (expressed as a percentage of total CVD deaths; denominator: all CVD deaths within each HIV-status and sex stratum) remained consistently higher among PLHIV than HIV-negative adults. In 2025, the proportion of CVD deaths attributable to elevated SBP was 73% (95% UI: 70-76%) among female PLHIV and 61% (95% UI: 58-64%) among male PLHIV, compared with 64% (95% UI: 61-66%) and 58% (95% UI: 55-60%) among HIV-negative females and males, respectively. By 2045, proportions increased modestly among PLHIV (female: 75% [95% UI: 72-78%]; male: 62% [95% UI: 59-65%]), while HIV-negative adults remained stable (Figure 4)

**Figure 4.**
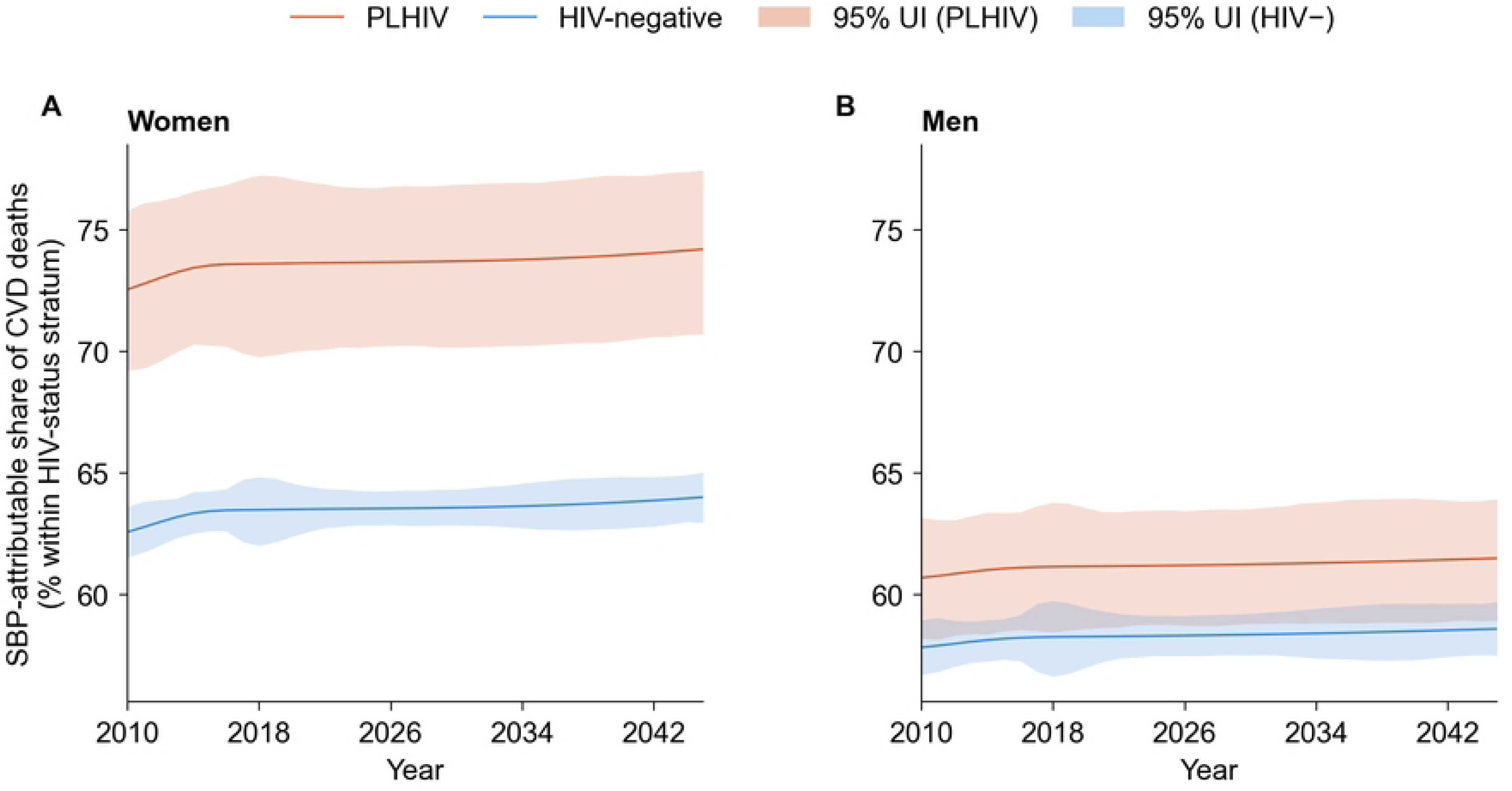
Proportion of CVD deaths attributable to high systolic blood pressure (SBP) under constant-RR assumptions. Estimated percentage of CVD deaths due to hypertension among PLHIV and HIV-negative adults, 2025-2045. The y-axis represents the proportion of total CVD deaths (denominator: all CVD deaths within each HIV-status and sex stratum) attributable to elevated SBP; it does not represent a population-level mortality rate. Under constant relative-risk assumptions, estimates change minimally over time, with consistently higher attributable fractions among PLHIV compared with HIV-negative adults.

### Scenario 4: Hypertension-associated CVD (RR increase post-DTG)

This scenario incorporated a stepwise increase in RR beginning in 2022, following Eswatini’s national rollout of DTG in 2021. Prior to 2021, the proportion of all CVD deaths attributable to high SBP were 73% (70-76%) among female PLHIV and 61% (58-64%) among male PLHIV, compared with 64% (61-66%) and 58% (55-60%) among their HIV-negative counterparts. Following the DTG rollout, a one-year-lagged transition in relative risk produced a sharp rise of approximately 12 percentage points in women (73 → 85%) and 9 points in men (61 → 70%) between 2021 and 2022. Thereafter, proportions remained largely stable through 2045. By 2045, estimates reached 85% (95% UI: 82-88%) among female PLHIV and 70% (95% UI: 67-73%) among male PLHIV, while corresponding values for HIV-negative adults remained unchanged (64% [95% UI 61-66%] in females; 58% [95% UI 55-60%] in males) (Figure 5).

**Figure 5.**
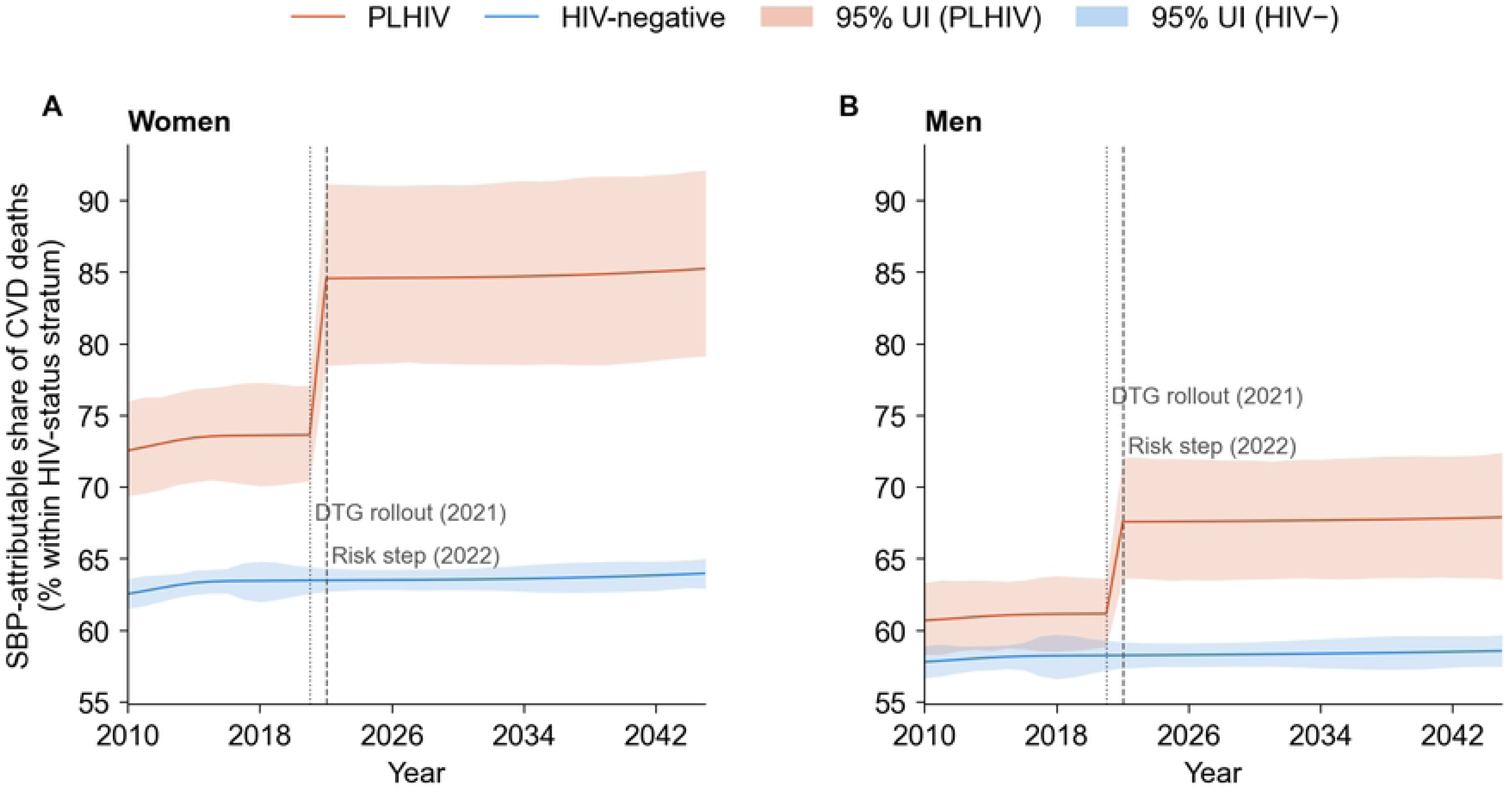
Hypertension-associated CVD mortality under post-DTG step-increase scenario. Proportion of all CVD deaths attributable to elevated systolic blood pressure (SBP) among PLHIV and HIV-negative adults. The denominator is total CVD deaths within each sex and HIV-status stratum. Relative risks increase over a one-year transition period following Eswatini’s 2021 dolutegravir rollout, reflecting staggered treatment uptake and delayed manifestation of DTG-associated metabolic effects.

### Scenario 5: Hypertension-Associated CVD (Gradual Ramp 2010–2045)

This scenario modeled a progressive, log-linear increase in HIV-associated RR from 2010 to 2045, representing cumulative influence of ART scale-up, survival-driven aging, and secular increases in hypertension and adiposity among PLHIV. At baseline (2010), the proportions of CVD deaths attributable to high SBP were similar between HIV-positive and HIV-negative adults, approximately 63-65% in women and 55-60% in men. By 2025, proportions had risen to 78% (95% UI 74-81%) among female PLHIV and 64% (95% UI 61-67%) among male PLHIV, compared with 66% (95% UI 63-68%) in HIV-negative females and 58% (95% UI 55-60%) in HIV-negative males. Notably, by this time, the burden among male PLHIV overtook that of HIV-negative females. Gradual divergence continued over the projection horizon, with 2045 estimates reaching 85% (95% UI 82-88%) among female PLHIV and 67% (95% UI 64-70%) among male PLHIV, compared with 60% and 55% among HIV-negative women and men, respectively (Figure 6)

**Figure 6.**
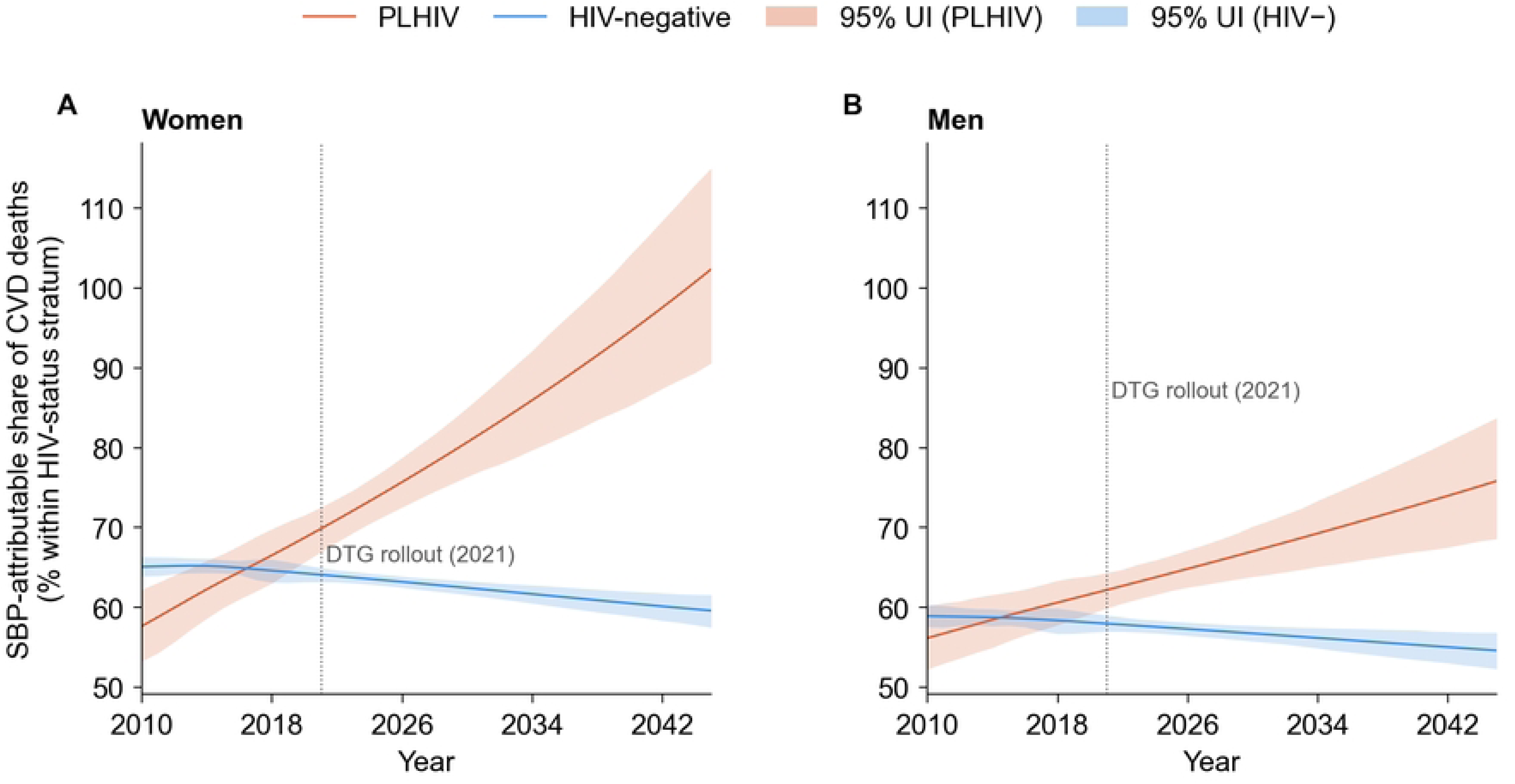
Gradual increase in hypertension-associated CVD burden. Relative risks increase log-linearly over time, reflecting cumulative effects of ART, aging, and secular hypertension trends. By 2045, female PLHIV reach 85% (95% UI 82-88%) and male PLHIV 67% (64-70%) of CVD deaths attributable to hypertension, widening the gap with HIV-negative adults.

### Sensitivity analyses

Projected CVD outcomes were robust to alternative extrapolation assumptions across all five scenarios. Replacing the generalized additive model with a logistic regression including a linear time trend produced qualitatively similar trajectories for CVD prevalence, CVD mortality and HTN-attributable CVD deaths and did not alter the direction or relative magnitude of differences by HIV status. Differences in absolute estimates were modest and did not affect conclusions regarding the projected CVD burden (Supplementary Figures S6, S8–S14). Results were similarly stable under probabilistic sampling of relative-risk assumptions and when evaluated over shorter analytic horizons (5- and 10-year periods) (Supplementary Table S4).

## Discussion

In this modeling analysis, we projected age-standardized cardiovascular disease (CVD) prevalence, annual CVD mortality risk, and the proportion of CVD mortality attributable to hypertension (HTN) among people living with HIV (PLHIV) in Eswatini through 2045. Across all scenarios, PLHIV consistently exhibited higher CVD prevalence and a greater proportion of CVD mortality attributable to HTN than HIV-negative adults, with particularly elevated risks among women. These findings align with a growing body of evidence showing that HIV infection and antiretroviral therapy (ART) contribute to increased cardiometabolic vulnerability through inflammatory, metabolic, and treatment-related pathways (53–55). While the absolute magnitude of differences varied across the four scenarios, the direction and persistence of disparities were robust, underscoring the need to integrate CVD prevention and management within HIV service delivery platforms.

Under the constant-risk scenarios, the gap in CVD prevalence between PLHIV and HIV-negative adults remained stable, suggesting that if current risk profiles do not worsen, the excess CVD burden in PLHIV may persist but not accelerate. However, the step-change and gradual-ramp scenarios indicated substantial potential for widening disparities, particularly under conditions reflecting the metabolic effects associated with dolutegravir (DTG) and ongoing increases in hypertension and adiposity (10,56,57). The stepwise increase in hypertension-attributable CVD following Eswatini’s 2021 DTG rollout mirrors cohort evidence showing clinically meaningful weight gain within 6–18 months of DTG initiation (9–11,56,58), which in the model increases HTN prevalence and, in turn, the proportion of CVD deaths attributable to high blood pressure. The progressive, log-linear divergence observed in the ramp scenario is consistent with longitudinal data demonstrating aging of the HIV-positive population, and secular increases in obesity and hypertension across the Southern African region (1,7,59), which are represented in the model through gradually increasing relative risks rather than explicit simulation of these processes. Together, these mechanisms suggest a sustained rise in CVD burden among PLHIV in the absence of proactive mitigation. In all projections, women with HIV exhibited the highest CVD burden, reflecting sex-specific vulnerability to ART-associated weight gain and higher baseline hypertension prevalence (28,29,43), which in the model translate into higher relative risks for hypertension-associated CVD among women compared with men.

Eswatini has already made substantial progress in integrating cardiometabolic risk screening and treatment within HIV care, including routine blood pressure screening, diabetes and lipid testing, lifestyle counseling, and, in some facilities, hypertension management using simplified clinical algorithms (15,60). Our results reinforce the importance of sustaining and scaling these integrated services as the population of people living with HIV ages and ART-related metabolic risks evolve. The modeling provides quantitative evidence of the potential rise in cardiovascular disease burden over the next two decades, highlighting the need for continued investment in integrated HIV-CVD prevention and treatment. These forecasts can help guide programmatic prioritization by identifying where scale-up of hypertension screening, CVD risk assessment, and treatment within ART clinics would yield the greatest long-term health impact. Nurse-led, decentralized hypertension management models are currently being piloted in Eswatini and have demonstrated feasibility; these approaches could be leveraged to achieve high coverage and blood pressure control even in resource-limited settings (60). The pronounced divergence observed among women underscores the need to strengthen sex-specific counseling and early intervention following DTG initiation, including proactive weight monitoring and management. More broadly, this analysis complements national efforts to institutionalize cardiometabolic surveillance within HIV cohorts and supports strategic planning and resource allocation under Eswatini’s National NCD and HIV strategies (15,60).

This study has several limitations. First, EMOD-HIV does not natively simulate cardiovascular disease (18), so CVD outcomes were incorporated through post-processing. As a result, potential feedback mechanisms such as whether CVD comorbidities may influence HIV-care retention, alter ART adherence, or increase mortality independently of HIV progression were not captured. While emerging observational studies suggest that chronic comorbid conditions may influence retention in care and adherence through increased care complexity and multimorbidity, the current evidence base remains insufficient to parameterize explicit CVD–HIV feedback mechanisms within the model (61,62). Second, CVD and hypertension prevalence inputs were drawn from secondary data sources (15,20,48), which are subject to sampling error, measurement variation, and methodological differences across survey rounds (21,46). However, most contributing surveys were nationally representative and based on probability samples aligned with Eswatini’s national census, supporting the validity of population-level estimates. Third, relative risk estimates were derived from published studies that varied in population characteristics, geographic setting, and follow-up duration. Although uncertainty was propagated using Monte Carlo simulation, structural sources of uncertainty such as non-linear changes in risk over time or unmeasured confounding in the source studies, were not fully captured. Fourth, the model assumed smooth temporal trends between observed survey years, which may not fully represent short-term fluctuations or external shocks, such as changes in treatment guidelines, health policy, or pandemics. Finally, long-term projections spanning two decades inherently carry uncertainty and should be interpreted as directional trends (supported by sensitivity analysis results) rather than precise predictions. In addition, the modeling assumes a simplified, mechanistic relationship in which hypertension increases CVD risk and CVD contributes to mortality through established risk-based pathways, without explicitly representing alternative causal structures, bidirectional effects, or heterogeneity in disease progression. These structural simplifications are appropriate for population-level planning but limit causal inference.

## Conclusion

Cardiovascular disease is likely to become an increasingly important contributor to morbidity and mortality among people living with HIV in Eswatini. Without targeted intervention, the excess burden borne by PLHIV, and particularly by women, is projected to persist and, under plausible metabolic and demographic trajectories, may widen substantially. Integrating hypertension screening, treatment, and CVD prevention strategies into ART delivery represents a critical opportunity to sustain the health gains achieved through HIV treatment scale-up and to advance long-term survival and well-being in high-burden settings.

## Data Availability

All data associated with the study are available within the manuscript and its Supporting Information.

## Consent for publication

Not applicable

## Availability of data and materials

All data used in this study were obtained from publicly available sources, including the Global Burden of Disease (GBD) study, World Health Organization STEPwise (STEPS) surveys, and published literature, as cited in the manuscript. Processed input data used in the analyses are provided in the Supplementary Methods, including GBD-derived CVD prevalence, CVD mortality, hypertension-attributable CVD mortality inputs, and relative risk estimates used in the scenario analyses. The EMOD-HIV model is publicly available as open-source software.

Analysis code is available from the corresponding author upon reasonable request.

## Competing interests

The authors declare that they have no competing interests.

## Funding

This work was supported by the National Heart, Lung, and Blood Institute (NHLBI) of the National Institutes of Health under Award Number 3R01HL169021-02S1 awarded to MPM. The content is solely the responsibility of the authors and does not necessarily represent the official views of the National Institutes of Health.

## Author contributions

MPM: Conceptualization, Methodology, Software, Validation, Formal analysis, Investigation, Data curation, Visualization, Writing – original draft, Writing – review & editing.

DTC: Software, Validation, Writing – review & editing.

KB: Writing – review & editing.

NY: Writing – review & editing.

AO: Writing – review & editing.

IP: Project administration, Writing – review & editing.

GF: Writing – review & editing.

CN: Resources, Writing – review & editing.

SGD: Writing – review & editing.

NG: Resources, Writing – review & editing.

AB: Conceptualization, Methodology, Supervision, Writing – review & editing

## Acknowledgements

The authors thank colleagues at the Department of Population Health, NYU Grossman School of Medicine, for valuable discussions during the development of this work.

## Supplementary Methods

**Figure S1. Observed versus modeled age- and sex-specific HIV prevalence in Eswatini across SHIMS survey years (2007, 2011, 2016, and 2021). Points and error bars represent empirical survey estimates; lines represent EMOD-HIV model outputs.** In particular, the model captured the temporal shift in peak HIV prevalence toward older age groups observed in successive survey rounds, reflecting changes in survival following ART scale-up. In addition to prevalence patterns, the model reproduced key demographic trends among people living with HIV. The mean age of the HIV-positive population increased steadily over time, rising from approximately 30 years in 2000 to 38–40 years by 2030 for both females and males (Figure S2).

**Figure S2. Observed and modeled mean age of people living with HIV (PLHIV) over time, by sex. Lines show model projections; markers indicate observed or survey-derived reference points where available.** These trends are consistent with observed demographic aging associated with sustained ART coverage and improved survival. Together, these calibration and validation results indicate that the model adequately represents both epidemiologic and demographic dynamics relevant to downstream projections of hypertension-associated cardiovascular disease burden.

**Figure S3. Age-standardized cardiovascular disease (CVD) prevalence by sex, Eswatini, 2010–2021.** Annual age-standardized CVD prevalence estimates (%) from the Global Burden of Disease (GBD) 2021 study for males, females, and both sexes combined. Lines represent point estimates and shaded bands represent 95% uncertainty intervals reconstructed from published GBD uncertainty bounds. CVD prevalence remained relatively stable over the study period, with consistently higher prevalence among males than females. Estimates are standardized to the GBD reference population.

**Figure S4. Age-standardized cardiovascular disease (CVD) mortality by sex, Eswatini, 2010–2021.** Annual age-standardized CVD mortality risk (%) derived from GBD 2021 mortality rates for males, females, and both sexes combined. Lines represent point estimates and shaded bands represent 95% uncertainty intervals. Mortality declined steadily throughout the study period for both sexes, while remaining consistently higher among males. Estimates are standardized to the GBD reference population and expressed as annual probability of dying from CVD.

**Figure S5. Age-standardized proportion of cardiovascular disease (CVD) deaths attributable to high systolic blood pressure (hypertension), by sex, Eswatini, 2010–2021.** Annual estimates of the fraction of CVD deaths attributable to elevated systolic blood pressure (%) obtained from the GBD 2021 study for males, females, and both sexes combined. Lines represent point estimates and shaded bands represent 95% uncertainty intervals. The hypertension-attributable fraction increased gradually over time and remained consistently higher among females than males. Estimates are age-standardized to the GBD reference population.

### Supplementary Note S1. Conversion of HRs to RRs

For HTN-associated CVD, published studies reported hazard ratios (HRs) rather than relative risks (RRs). To integrate these estimates into our modeling framework, we converted HRs into RRs using the following steps:

#### Step 1. Baseline mortality rates from GBD 2021

We obtained age-standardized CVD mortality rates attributable to high systolic blood pressure (SBP) for Eswatini from the Global Burden of Disease (GBD) 2021 study. These rates were reported as deaths per 100,000 person-years:

Males: 392.4 | Females: 325.6 | Both sexes: 360.7

#### Step 2. Conversion to annual hazard rate (***r*_0_**)

We first expressed these rates on a per person-year basis by dividing by 100,000:

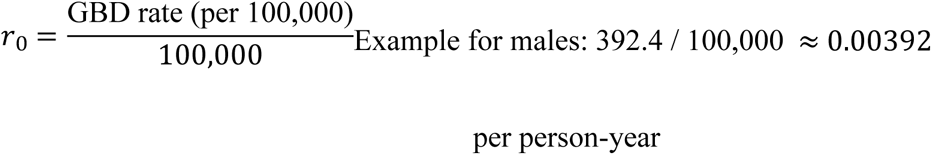

#### Step 3. Conversion to cumulative baseline risk (***R*_0_**)

Because GBD reports rates (not risks), we converted *r*_0_into a cumulative risk over the approximate follow-up period of the HR studies (20 years), assuming exponential survival (Greenland, 1987): *R*_0_ = 1 ― *e*^―*r*0⋅*t*^

Example for males: *R*_0_ = 1 ― *e*^―(0.00392×20)^ ≈ 0.0392 (or 3.92% over 20 years)

#### Step 4. Conversion of HR to RR

We then applied the Zhang & Yu (1998) approximation, which adjusts HRs for baseline risk:

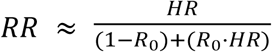

This ensures the relative risk reflects the observed baseline incidence in the unexposed group (HIV-negative adults).

#### Step 5. Application to sex-specific estimates

This procedure was repeated for males, females, and all-sex categories using their respective baseline risks. Converted RRs (with 95% CIs) are presented in Table S1.

**Table S1.** Relative risks (RRs) of HTN-associated CVD in PLHIV versus HIV-negative adults, converted from hazard ratios (HRs) using GBD baseline risks (Eswatini, 2021) by Mohammed Siddiqui et al., 2023.

| HIV status | Sex | HR (95% CI) | Baseline risk P0 | Converted RR (95% CI) |
| --- | --- | --- | --- | --- |
| HIV-negative | Male | 1.51 (1.38–1.64) | 0.00392 | 1.50 (1.36–1.63) |
|  | Female | 2.01 (1.44–2.82) | 0.00326 | 1.99 (1.42–2.79) |
| HIV-positive | Male | 1.67 (1.46–1.92) | 0.00392 | 1.66 (1.45–1.90) |
|  | Female | 2.25 (1.76–2.88) | 0.00326 | 2.23 (1.74–2.85) |
| HIV-negative | All sexes | 1.56 (1.44–1.69) | 0.00361 | 1.55 (1.43–1.68) |
| HIV-positive | All sexes | 1.73 (1.52–1.96) | 0.00361 | 1.72 (1.51–1.95) |

To obtain relative risks of HTN-associated CVD in PLHIV versus HIV-negative adults, the converted RRs for HIV-positive individuals were divided by the corresponding converted RRs for HIV-negative individuals (Table S2).

**Table S2.** Relative risks (RRs) of HTN-associated CVD in PLHIV versus HIV-negative adults, obtained by dividing converted RRs for HIV-positive individuals by corresponding values for HIV-negative individuals.

| Sex | Converted RR (95% CI) | RR (95% CI) |
| --- | --- | --- |
| Male | 1.66 (1.45–1.90) | 1.11 (1.06 - 1.16) |
| Female | 2.23 (1.74–2.85) | 1.12 (1.02 - 1.22) |
| All sexes | 1.72 (1.51–1.95) | 1.11 (1.06 - 1.16) |

**Table S3.** Relative risks (RRs) used in CVD forecasting scenarios.

| Scenario | Outcome | Time pattern | Women (95% UI) | Men (95% UI) | Both sexes (95% UI) | Sources |
| --- | --- | --- | --- | --- | --- | --- |
| **Scenario 1:**<br>General CVD<br>(constant RR) | All CVD | Constant,<br>2010–2045 | — | — | 1.60 (1.30–2.00) | Published comparative RR for PLHIV vs HIV-negative adults (31) |
| **Scenario 2:**<br>General CVD mortality (constant RR) | All CVD | Constant, 2010–2045 | — | — | 1.60 (1.30–2.00) | Published comparative RR for PLHIV vs HIV-negative adults (31) |
| **Scenario 3:**<br>HTN-associated CVD (constant RR) | HTN-CVD | Constant, 2010–2045 | 1.16 (1.10–1.22) | 1.05 (1.00–1.10) | 1.10 (1.05–1.15) | Siddiqui et al.; HRs converted to RRs using Eq. 1 (30,31) |
| **Scenario 4:**<br>HTN-associated CVD (post-DTG step increase) | HTN-CVD | Constant through 2021 (same as Scenario 3); step increase in 2022; constant thereafter | 1.16 → 1.40 (1.25–1.50) | 1.05 → 1.20 (1.10–1.30) | 1.10 → 1.29 (1.18–1.40) | DTG-associated weight gain translated to BMI-CVD risk; ~1-year lag post-DTG rollout (8–10, 14, 44–46) |
| **Scenario 5:**<br>HTN-associated CVD (gradual ramp) | HTN-CVD | Log-linear increase, 2010–2045 | 1.05 → 1.43 (1.30–1.55) | 1.05 → 1.25 (1.15–1.35) | 1.10 → 1.33 (1.22–1.45) | ART-era anchoring + DTG, aging, secular adiposity/HTN trends (1,3,14,47,48) |

Notes: Sex-specific RRs were used for sex-stratified analyses (Scenarios 3–5); pooled "both sexes" RRs were used in analyses not stratified by sex (Scenarios 1–2). In Scenario 4, pre-2022 RRs are identical to Scenario 3, with a step increase applied beginning in 2022 following Eswatini's national DTG rollout. Full derivations for Scenario 4 step changes and Scenario 5 target values are provided in Supplementary Note S2 and S3.
RR = relative risk; UI = uncertainty interval; HTN = hypertension; CVD = cardiovascular disease; PLHIV = people living with HIV; DTG = dolutegravir; BMI = body mass index.

**Table S4.** Projected CVD outcomes by HIV status across analytic horizons — Sensitivity Analysis 2.

| Scenario | Analytic horizon | Year | PLHIV (%) | HIV-negative (%) | Difference (pp) |
| --- | --- | --- | --- | --- | --- |
| **Scenario 1: General CVD prevalence (both sexes)** |  |  |  |  |  |
| *Metric: CVD prevalence (%)* |  |  |  |  |  |
|  | 5-year (2025–2030) | 2030 | 11.21 | 6.90 | **4.31** |
|  | 10-year (2025–2035) | 2035 | 11.21 | 6.90 | **4.31** |
|  | 20-year (2025–2045) | 2045 | 11.23 | 6.91 | **4.32** |
| **Scenario 2: General CVD mortality (both sexes)** |  |  |  |  |  |
| *Metric: CVD mortality rate (%)* |  |  |  |  |  |
|  | 5-year (2025–2030) | 2030 | 0.59 | 0.36 | **0.23** |
|  | 10-year (2025–2035) | 2035 | 0.57 | 0.35 | **0.22** |
|  | 20-year (2025–2045) | 2045 | 0.54 | 0.33 | **0.21** |
| **Scenario 3: HTN-attributable CVD deaths, constant RR (both sexes)** |  |  |  |  |  |
| *Metric: SBP-attributable CVD deaths (% within HIV-status stratum)* |  |  |  |  |  |
|  | 5-year (2025–2030) | 2030 | 70.43 | 60.75 | **9.68** |
|  | 10-year (2025–2035) | 2035 | 70.54 | 60.85 | **9.70** |
|  | 20-year (2025–2045) | 2045 | 70.91 | 61.17 | **9.75** |
| **Scenario 4: HTN-attributable CVD deaths, step RR post-DTG 2022 (both sexes)** |  |  |  |  |  |
| *Metric: SBP-attributable CVD deaths (% within HIV-status stratum)* |  |  |  |  |  |
|  | 5-year (2025–2030) | 2030 | 80.29 | 58.65 | **21.64** |
|  | 10-year (2025–2035) | 2035 | 80.40 | 58.73 | **21.67** |
|  | 20-year (2025–2045) | 2045 | 80.84 | 59.05 | **21.79** |
| **Scenario 5: HTN-attributable CVD deaths, gradual ramp RR (both sexes)** |  |  |  |  |  |
| *Metric: SBP-attributable CVD deaths (% within HIV-status stratum)* |  |  |  |  |  |
|  | 5-year (2025–2030) | 2030 | 76.74 | 59.41 | **17.34** |
|  | 10-year (2025–2035) | 2035 | 82.70 | 58.24 | **24.46** |
|  | 20-year (2025–2045) | 2045 | 95.85 | 55.85 | **40.00** |

*pp = percentage points. Projections use a logistic generalized additive model (GAM) fitted to GBD 2010–2021 data and extrapolated forward. Horizons are anchored to 2025 as the baseline projection year. PLHIV = people living with HIV. RR = relative risk; DTG = dolutegravir; SBP = systolic blood pressure.*

### Supplementary Note S2. How DTG-associated weight gain was translated into CVD relative risks

#### Step 1. Excess weight gain with DTG

Randomized trials show greater weight gain with dolutegravir (DTG) compared with efavirenz (EFV): participants on DTG gained approximately 3.5 kg more than those on EFV at 192 weeks, and women receiving DTG/TAF gained about 4.8 kg more than those on EFV at 96 weeks (29).

#### Step 2. Converting weight gain to BMI increase

Excess weight gain (Δ*W*) was converted to a change in body mass index (BMI) using: 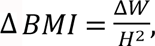 where *H* is height (m). Adult heights were taken from the WHO STEPS Eswatini survey (mean 1.58 m for women and 1.68 m for men) (44). Using these values, a 3.5 kg excess gain corresponds to Δ*BMI* ≈ 1.24 *for men and* 1.40 *for women*, while a 4.8 kg gain corresponds to Δ*BMI* ≈ 1.70 for men and 1.92 for women.

#### Step 3. Mapping ΔBMI to CVD relative risk

From prior studies (46,60), vascular mortality risk increases by about 40% for every 5 kg/m² increase in BMI. To translate this into smaller BMI changes, we assumed the relationship is log-linear, that is, each 1-unit increase in BMI multiplies risk by the same factor.

- **Step 1:** From the published finding (46): +5 BMI → relative risk (RR) = 1.40.
- **Step 2:** We spread this evenly across 5 units by taking the fifth root of 1.40: *R R*_per 1 BMI_ = 1.40^1/5^ ≈ 1.069, meaning each 1-unit increase in BMI raises CVD risk by ∼6.9%.
- **Step 3:** For any change in BMI (Δ*BMI*), we raised 1.069 to the power of Δ*BMI*: *R R*_multiplier_ = 1.069^Δ*BMI*^. Applying these to our sex-specific Δ*BMI* values yields:

- 3.5 kg excess gain

- Men: Δ*BMI* = 1.24⇒*RR* ≈ 1.086 (+8.6%)
- Women: Δ*BMI* = 1.40⇒*RR* ≈ 1.098 (+9.8%)
- 4.8 kg excess gain

- Men: Δ*BMI* = 1.70⇒*RR* ≈ 1.120 (+12.0%)
- Women: Δ*BMI* = 1.92⇒*RR* ≈ 1.137 (+13.7%)

This corresponds ∼9–10% higher CVD risk for 3.5 kg excess gain and ∼12–14% for a 4.8 kg gain, with larger effects in women than men. In our scenario analyses, we modeled slightly larger step increases (∼18% in women and ∼14% in men) to account for additional DTG-associated metabolic effects beyond BMI alone.

### Supplementary Note S3. Decomposition of RR trajectories in the gradual-ramp scenario (scenario 5)

Scenario 5 represents a gradual, long-term increase in hypertension-associated cardiovascular disease (HTN-CVD) risk among PLHIV relative to HIV-negative adults between 2010 and 2045. Rather than assuming an abrupt change in RR, this scenario reflects the combined influence of several slow-moving, compounding drivers that elevate relative risk over time. These processes were represented as multiplicative calibration components, including dolutegravir (DTG)-associated metabolic effects, survival-driven aging of the PLHIV population, and secular increases in adiposity and hypertension in the background population, each contributing to the widening PLHIV–HIV-negative gap.

These multipliers were not modeled as independent causal estimates and were not used as uncertainty-analysis ranges. Instead, they were specified as transparent, approximate calibration components chosen to generate plausible 2045 RRs consistent with the hypothesized epidemiologic trajectory in each stratum. Table S5A summarizes the conceptual interpretation and empirical motivation for each calibration component, whereas Table S5B reports the exact sex-specific values used to ensure that the product of the baseline RR and the component multipliers matched the target 2045 RR in each stratum.

#### Framework

For each stratum, the target RR in 2045 was defined as:

^*RR*^2045 ^= *RR*^2010 ^× *M*^DTG ^× *M*^Aging ^× *M*^Secular

where:

- *RR*_2010_ is the baseline RR in 2010;
- *M*_DTG_ is the multiplier representing gradual DTG-associated metabolic effects;
- *M*_Aging_ is the multiplier representing survival-driven aging of the PLHIV population; and
- *M*_Secular_is the multiplier representing secular increases in adiposity and hypertension in the background population.

Baseline RRs in 2010 were set to 1.05 for women, 1.05 for men, and 1.10 for the combined-sex analysis. Target RRs in 2045 were set to 1.43 for women, 1.25 for men, and 1.33 for the combined-sex analysis.

#### Interpretation and empirical motivation of calibration components

The three calibration components were chosen to reflect plausible long-term drivers of increasing HTN-CVD risk among PLHIV. As shown in Table S5A, *M*_DTG_ was motivated by evidence on DTG-associated weight gain and metabolic effects, *M*_Aging_ by evidence that ART scale-up extends survival and shifts the PLHIV population toward older ages, and *M*_Secular_by evidence of rising obesity and hypertension in Eswatini and the broader southern African region.

**Table S5A.** Literature-informed interpretation of calibration components in Scenario 5 (Gradual Ramp)

| Component | Description | Empirical basis / justification | Approximate cumulative contribution by 2045 |
| --- | --- | --- | --- |
| $M_{DTG}$ | Gradual metabolic and weight-gain effects associated with dolutegravir (DTG), modeled as a smooth ramp beginning in 2021 | ADVANCE and NAMSAL trials; BMI-to-CVD conversion informed by Prospective Studies Collaboration and related literature | Approximately 10% for men and 15% for women |
| $M_{Aging}$ | Increased exposure to CVD risk factors as ART scale-up extends survival and shifts the PLHIV population toward older ages | Nakagawa et al., 2018; Harris et al., 2021; UNAIDS, 2013 | Approximately 5%–10% |
| $M_{Secular}$ | Background rise in obesity and hypertension in Eswatini and the broader southern African region | WHO STEPS Eswatini, 2024; Gona et al., 2021; Barry et al., 2025; WHO AFRO, 2024 | Approximately 10%–15% |

**Note:** Approximate contributions represent literature-informed conceptual magnitudes by 2045 and are not uncertainty-analysis ranges or independently estimated causal effects.

The approximate cumulative contributions shown in Table S5A are intended to convey the conceptual magnitude of each component by 2045; they are not uncertainty bounds and were not applied directly as ranges in the model.

#### Exact calibration values used to match the 2045 targets

The exact calibration values used in the model are shown in Table S5B.

**Table S5B.**
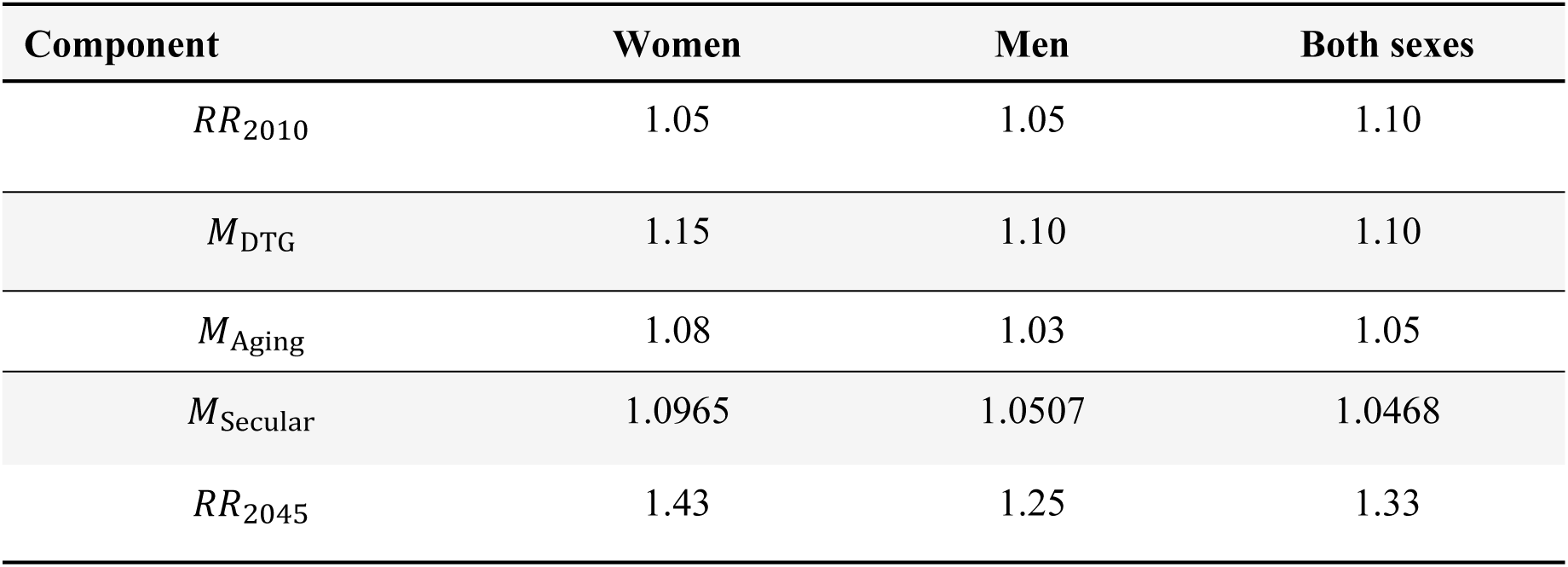
Exact calibration values used to match the target 2045 relative risks in Scenario 5 (Gradual Ramp)

These values were selected jointly within each stratum so that the product *RR*_2045_ = *RR*_2010_ × *M*_DTG_ × *M*_Aging_ × *M*_Secular_ equaled the target 2045 RR exactly. These calibration values satisfy the target RRs exactly:

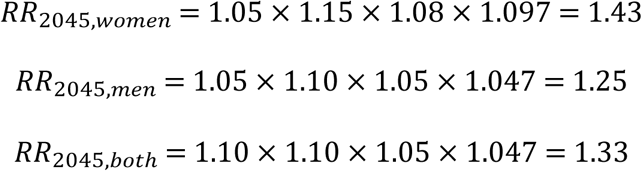

#### Interpretation of component multipliers

The component multipliers should be interpreted as cumulative calibration factors operating over the full 2010-2045 period. For example *M*_Aging_1.08 for women indicates an assumed 8% cumulative increase in RR by 2045 attributable to survival-driven aging of the PLHIV population, relative to the 2010 baseline. These values do not represent annual increases. Rather, they summarize the total contribution of each component to the 2045 RR endpoint.

#### Temporal implementation

After specifying *RR*_2010_and *RR*_2045_, the year-specific RR trajectory was modeled as a smooth log-linear increase from 2010 to 2045:

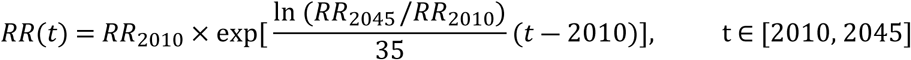

This formulation yields a continuous increase in RR over time, consistent with gradual epidemiologic change rather than an abrupt intervention-induced shift (Figure S3).

**Figure S6. Relative risk trajectories applied to PLHIV versus HIV-negative adults – HTN-attributable CVD Deaths, Scenarios 3-5 (2010 – 2045)**

## Steps: Estimating and forecasting CVD outcomes

**Step 1:** Decomposing total population CVD estimates into HIV-status strata.

General population age-standardized CVD outcomes were obtained from GBD 2021 annual estimates for Eswatini (2010–2021; Supplementary Table S1).

Because GBD provides total population estimates rather than HIV-stratified values, CVD outcomes among HIV-negative adults were derived using a conditional probability decomposition. The total population CVD outcome can be expressed as:

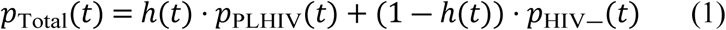

where ℎ(*t*)is the HIV prevalence in the general population at time *t*, derived from calibrated EMOD-HIV model outputs. Applying the scenario-specific relative risk RR(*t*) - defined as the ratio of the CVD outcome in PLHIV relative to HIV-negative adults - gives *p*_PLHIV_(*t*) = RR (*t*) × *p*_HIV―_(*t*). Substituting and solving for *p*_HIV―_(*t*):

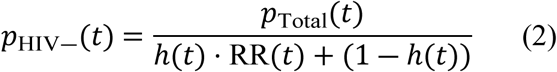

CVD outcomes among PLHIV were then recovered as:

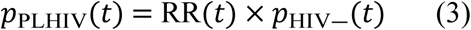

These reconstructed time series (2010–2021) represent the empirical baseline derived from GBD, EMOD-HIV, and published RR estimates (Supplementary Table S3). They served as the basis for forecasting future trends, with uncertainty propagated through Equations (2) and (3) as described in the Uncertainty Analysis section.

**Step 2:** Forecasting HIV-negative CVD outcomes using a generalized additive model.

To project CVD prevalence among HIV-negative adults beyond 2021, we fitted a generalized additive model (GAM) with a logistic link function to the HIV-negative series derived in Step 1:

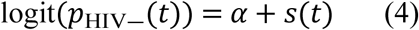

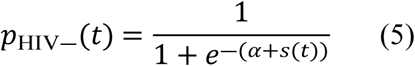

where *p*_HIV―_(*t*)is the predicted HIV-negative CVD outcome in year *t*, *α*is the intercept, and *s*(*t*) is a smooth function of time estimated using penalized splines. The logistic link constrains predicted values between 0 and 1, while penalized splines capture smooth, nonlinear temporal trends. The fitted model was extrapolated from 2022 to 2045, assuming that long-term changes continue smoothly along the observed temporal trajectory. Logistic regression with a linear time trend was used as a sensitivity analysis (38, 39).

**Step 3:** Forecasting CVD outcomes among PLHIV.

Forecasted CVD outcomes among PLHIV were derived by applying the scenario-specific relative risk RR(*t*)to the corresponding HIV-negative forecast from Step 2 for each year:

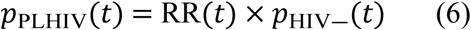

This formulation maintains internal consistency across scenarios and time periods, ensuring that stratum-specific estimates remain anchored to the HIV-negative baseline and reflect only the scenario-specific excess CVD risk among PLHIV.

**Figure S7. Analytical workflow for projecting cardiovascular disease burden among adults in Eswatini, 2025–2045.** Workflow integrating EMOD-HIV outputs, GBD 2021 inputs, published relative risks, GAM-based forecasting, and Monte Carlo uncertainty quantification to project five CVD scenarios through 2045. Outcomes for PLHIV and HIV-negative adults were estimated using RR-based decomposition of GBD-derived population measures.

**Figure S8. Modeled cardiovascular disease (CVD) prevalence by HIV status and sex.** Age-standardized CVD prevalence (%) with 95% uncertainty intervals under a constant relative-risk (RR = 1.6[1.3-2.0]) assumption. Male PLHIV exhibited the highest prevalence (∼11.7%, 95% UI: 9.5–13.5% in 2025), followed by female PLHIV (∼10.8%, 95% UI: 9.0–12.5%), male HIV-negative adults (∼7.2%, 95% UI: 6.6–7.8%), and female HIV-negative adults (∼6.6%, 95% UI: 6.2–7.2%), with all differences remaining stable and non-overlapping through 2045.

**Note:** Projected CVD prevalence was higher among male PLHIV than female PLHIV, a pattern driven by the decomposition method: because HIV prevalence is lower among men than women in Eswatini, the relative-risk adjustment concentrates excess CVD burden across a smaller PLHIV denominator, yielding higher estimated prevalence per PLHIV individual. This reflects a mathematical consequence of the decomposition framework rather than a biological difference in CVD risk by sex, and should be interpreted accordingly.

**Figure S9. Modeled general CVD mortality risk by HIV status. Age-standardized annual probability of dying from CVD (%) with 95% uncertainty intervals under a constant relative-risk (RR = 1.6 [1.3–2.0]) assumption.** PLHIV consistently exhibited higher CVD mortality risk than HIV-negative adults across both sexes throughout the projection horizon, with a modest declining trend reflecting GBD-derived mortality inputs.

**Figure S10. Sensitivity analysis: GAM versus linear time trend extrapolation — Scenario 1 (General CVD Prevalence, Constant RR).** Age-standardized CVD prevalence (%) among PLHIV and HIV-negative adults projected from 2010 to 2045 using a generalized additive model (GAM; left panel) and a logistic regression with a linear time trend (right panel). Both sexes combined; RR = 1.6 (95% CI: 1.3–2.0). Shaded ribbons represent 95% uncertainty intervals.

**Figure S11. Sensitivity analysis: GAM versus linear time trend extrapolation — Scenario 2 (General CVD Mortality, Constant RR).** Annual CVD death probability (%) among PLHIV and HIV-negative adults projected from 2010 to 2045 using a generalized additive model (GAM; left panel) and a logistic regression with a linear time trend (right panel). Both sexes combined; RR = 1.6 (95% CI: 1.3–2.0). Shaded ribbons represent 95% uncertainty intervals.

**Figure S12. Sensitivity analysis: GAM versus linear time trend extrapolation — Scenario 3 (HTN-Attributable CVD Deaths, Constant RR).** SBP-attributable share of CVD deaths (% within HIV-status stratum) among PLHIV and HIV-negative adults projected from 2010 to 2045 using a generalized additive model (GAM; left panel) and a logistic regression with a linear time trend (right panel). Both sexes combined; Shaded ribbons represent 95% uncertainty intervals.

**Figure S13. Sensitivity analysis: GAM versus linear time trend extrapolation — Scenario 4 (HTN-Attributable CVD Deaths, Post-DTG Step Increase).** SBP-attributable share of CVD deaths (% within HIV-status stratum) among PLHIV and HIV-negative adults projected from 2010 to 2045 using a generalized additive model (GAM; left panel) and a logistic regression with a linear time trend (right panel). Step increase in RR applied to PLHIV from 2022 following Eswatini’s national dolutegravir rollout; HIV-negative adults held constant. Both sexes combined; sex-specific RRs applied. Shaded ribbons represent 95% uncertainty intervals.

**Figure S14. Sensitivity analysis: GAM versus linear time trend extrapolation — Scenario 5 (HTN-Attributable CVD Deaths, Gradual Ramp).** SBP-attributable share of CVD deaths (% within HIV-status stratum) among PLHIV and HIV-negative adults projected from 2010 to 2045 using a generalized additive model (GAM; left panel) and a logistic regression with a linear time trend (right panel). Log-linear RR increase from 2010 anchored to early-ART-era estimates, reaching target values by 2045; applied to PLHIV only. Both sexes combined; sex-specific RRs applied. Shaded ribbons represent 95% uncertainty intervals.

## Notes

### Competing Interest Statement

The authors have declared no competing interest.

